# The Use and Safety Risk of Repurposed Drugs for COVID-19 patients: Lessons Learned Utilizing the Food and Drug Administration’s Adverse Event Reporting System

**DOI:** 10.1101/2022.12.10.22283298

**Authors:** Deemah S. Alsuhaibani, Heba H. Edrees, Thamir M Alshammari

**Author notes:** **Corresponding Author Thamir M. Alshammari, Ph.D, M.S, RPh, FISoP, CCPDC**, Associate professor of pharmacy practice, Senior Researcher, Medication Safety Research Chair, King Saud University, Riyadh, Saudi Arabia, College of Pharmacy, Almaarefa University, Riyadh, Saudi Arabia, Pharmacoepidemiology and Pharmacovigilance Consultant, P.O. Box 2457, Riyadh 11451.

## Abstract

**Objectives:** This study was designed to assess the disproportionality analyses of adverse drug reactions (ADRs) related to hydroxychloroquine and remdesivir and how ADR reporting fluctuated during the COVID-19 pandemic.

**Methods:** A retrospective observational study was conducted utilizing the Food and Drug Administration’s Adverse Event Reporting System (FAERS) data between 2019 and 2021. The study was conducted in two phases. In the first phase, all reports associated with the drugs of interest were evaluated to assess all related adverse drug reactions. In the second phase, specific outcomes of interest (i.e., QT prolongation and renal and hepatic events) were determined to study their association with the drugs of interest. A descriptive analysis was conducted for all adverse reactions related to the drugs being studied. In addition, disproportionality analyses were conducted to compute the reporting odds ratio, the proportional reporting ratio, the information component, and the empirical Bayes geometric mean. All analyses were conducted using RStudio.

**Results:** A total of 9,443 ADR reports related to hydroxychloroquine; 6,160 (71.49) patients were female, and higher percentage of patients of both sexes were over the age of 65 years. QT prolongation (1.48%), pain (1.38%), and arthralgia (1.25%) were most frequently reported ADRs during the COVID-19 pandemic. The association of QT prolongation with use of hydroxychloroquine was statistically significant (ROR 47.28 [95% CI 35.95-62.18]; PRR 42.41 [95% CI 32.25-55.78]; EBGM 16.08; IC 4.95) compared with fluoroquinolone. The outcome was serious medical events in 48.01% of ADR reports; 27.42% required hospitalization and 8.61% resulted in death. Of 6,673 ADR reports related to remdesivir, 3,928 (61.13%) patients were male. During 2020, the top three ADR reports were elevated liver function tests (17.26%), acute kidney injury (5.95%) and death (2.84%). Additionally, 42.71% of ADR reports indicated serious medical events; 19.69% resulted in death and 11.71% indicated hospitalization. The ROR and PRR of hepatic and renal events associated with remdesivir were statistically significant, (4.81 [95% CI 4.46-5.19] and 2.96 [95% CI 2.66-3.29], respectively.

**Conclusion:** Our study showed that several serious ADRs were reported with the use of hydroxychloroquine, which resulted in hospitalization and death. Trends with the use of remdesivir were similar, but to a lesser extent. Therefore, this study showed us that off-label use should be based on thorough evidence-based evaluation.

## Introduction

The Coronavirus Disease 2019 (COVID-19) emerged in late 2019 and, to date, there have been more than 612 million cumulative cases and more than 6.5 million cumulative deaths reported worldwide. (1) There is still extensive research being conducted on COVID-19, especially regarding the pathophysiology and drug therapy. Since the beginning of the pandemic, the safety and efficacy of several drug therapies linked to COVID-19 have been questionable. Several existing drugs have been considered potential therapies for COVID-19, including chloroquine, hydroxychloroquine, and remdesivir. (2)

Chloroquine and hydroxychloroquine showed in vitro viral inhibition and were given emergency approval by the United States Food and Drug Administration (FDA) and the European Medicines Agency for use in hospitalized patients with COVID-19. (3-6) However, several randomized clinical trials showed these drugs were not effective in treating COVID-19 or for post-exposure prophylaxis. Although hydroxychloroquine reduced the length of hospitalization, it was associated with a higher COVID-19 mortality risk and adverse effects including QT prolongation, hepatotoxicity, low lymphocyte level, and thrombocytopenia. (7-9) As a result, the FDA revoked its emergency approval, and these drugs are no longer considered useful for treating COVID-19. Remdesivir also received FDA emergency use authorization in May 2020 and FDA approval in October 2020. In patients with severe COVID-19, remdesivir prominently shortened the time of clinical improvement and reduced mortality. (10) However, a clinical trial and one small clinical study showed that several adverse drug reactions (ADRs) were reported with remdesivir, including hepatotoxicity, nephrotoxicity, gastrointestinal discomfort, hypotension, and respiratory toxicity. (11, 12)

With the constant changes and development of treatment options for COVID-19, it is important to report ADRs. One useful tool for reporting ADRs is the FDA’s Adverse Event Reporting System (FAERS). FAERS is a database that helps with post-marketing safety surveillance for drugs and therapeutic biologics. Reports can be submitted by health care professionals, consumers, and manufacturers. The objectives of this study were to identify the disproportionality analyses of the events associated with hydroxychloroquine and remdesivir and to determine how ADR reporting for hydroxychloroquine and remdesivir changed from 2019 to 2021.

## Methods

### Study design

This was a retrospective observational study using the FAERS. It was conducted utilizing FAERS data between 2019 and 2021. FAERS was established in 1969 and includes adverse events and medication error reports related to regular medicinal products and biologics. Access to this data is free and is available on the FDA website. (13)

### Data sources

FAERS publishes its files quarterly, and the files are arranged into 7 categories: demographics, drugs, indications, outcomes, reactions, reporting source, and therapies. These files include detailed information related to each topic. All files include one unique variable to facilitate merging these files whenever needed. This variable is called the “primaryid” in files published after 2013. For those published before 2013, this unique variable was called “individual safety report.” FAERS accept reports from manufacturers, health care professionals, and patients/consumers. Reporting ADRs is mandatory for manufacturers, while it is voluntary for health care professionals and patients/consumers. (14)

### Outcomes of interest

The aim of this study was to analyze the adverse event reports of two medications, hydroxychloroquine and remdesivir, most commonly used for COVID-19. The ADR reports were from 2019, 2020, and 2021. The analyzed drugs’ role codes were “primary and secondary suspected drugs” and “concomitant drugs” to ensure all related adverse events were included.

The association between specific adverse events with the drugs of interest was assessed based on QT prolongation events for patients who used hydroxychloroquine, as this is a serious adverse event. The outcomes of interest for remdesivir were renal and hepatic events. Using preferred terms, the terms “QT” and “prolongation” were used to find QT events, while “renal,” “hepatic,” “kidney,” and “liver” were used to find renal and hepatic events. Here, the drugs analyzed needed to be the primary drug related to the adverse event. Duplicate reports were excluded by checking the primary suspected and the case number.

### Drugs of interest

Two drugs were included in the study. Hydroxychloroquine was one of the most commonly used medications for COVID-19, and its effectiveness was debated. The other drug, remdesivir, was the only drug approved by the United States FDA at the peak of the pandemic to treat patients with COVID-19. (13, 15) The reason for analyzing hydroxychloroquine is because of its increased use, but lack of approval, to treat COVID-19; this study was intended to assess the association between the signal of QT prolongation and use of hydroxychloroquine. For comparison purposes, the same signal was evaluated with fluoroquinolone medications use because of the known risk of QT prolongation with fluoroquinolone use. (16) The fluoroquinolone medications included were ciprofloxacin, levofloxacin, moxifloxacin, and norfloxacin. Drug indications were assessed to make sure that only indications of COVID-19 and related keywords such as coronavirus, corona, and COVID were included. All other indications for these medications were excluded.

### Statistical analysis

Two types of analyses were conducted in this study. First, descriptive statistics were used for different variables, including demographic and medical data. The data was presented for each year individually and for all years combined. Second, disproportionality analyses were computed, including reporting odds ratio (ROR), proportional reporting ratio (PRR), information component (IC), and empirical Bayes geometric mean (EBGM). These analyses were used to assess the association between the signal of QT prolongation development with use of hydroxychloroquine and between the signal of renal and hepatic development with the use of remdesivir.

A case–non-case methodology was used to evaluate these associations. A contingency table including 4 components was used to calculate disproportionality metrics (i.e., ROR, PRR, IC, and EBGM), and these 4 components (cells) were labeled “a,” “b,” “c,” and “d.” Cell “a” represents number of reports of cases (outcome of interest) for the studied drugs, while “b” is the number of reports of non-cases for those drugs. “C” represents the number of reported cases for other medications, and “d” represents the number of reports of non-cases for all other medications. The statistical equations for calculating these 4 metrics are explained elsewhere (17). All statistical analyses were conducted using R (version 4.1.3) and RStudio (2022.7.1.554) software.

## Results

From 2019 to 2021, there were 9,443 hydroxychloroquine-related ADR reports and 6,673 remdesivir-related reports submitted to FAERS. We analyzed the demographical data, indications, and outcomes related to each ADR reported to FAERS during the study period.

The characteristics of patients with ADRs are reported in Table 1. Overall, for hydroxychloroquine, females experienced a higher incidence of ADRs (more than twice as often as males), with the majority of individuals > 65 years (2,917, 30.89%).

**Table 1:** Characteristics of patients with ADRs related to hydroxychloroquine and remdesivir.

| Characteristic | Hydroxychloroquine<br>(n = 9,443)<br>n (%) | Remdesivir<br>(n = 6,673)<br>n (%) |
| --- | --- | --- |
| Male | 2,453 (28.47) | 3,928 (61.13) |
| Female | 6,160 (71.49) | 2,496 (38.84) |
| Age group: |  |  |
| < 30 years | 594 (6.29) | 436 (6.53) |
| 31-50 years | 1,675 (17.74) | 1,098 (16.45) |
| 51-60 years | 1,847 (19.56) | 1,020 (15.29) |
| 61-65 years | 925 (9.8) | 602 (9.02) |
| > 65 years | 2,917 (30.89) | 2,949 (44.19) |
| Source of the report | Consumer* | Pharmacist* |
| Reporting country | US, Canada, and France* | US, Japan, and Italy* |

**Table 1:** Outcomes related to hydroxychloroquine and remdesivir.

| Hydroxychloroquine |  |  |  |
| --- | --- | --- | --- |
| Outcomes | 2019<br>(%) | 2020<br>(%) | 2021<br>(%) |
| Not available | 55.95 | 10.32 | - |
| Other serious (important medical event) | 29.52 | 48.01 | 61.75 |
| Required intervention to prevent permanent impairment/damage | 0.03 | 0.02 | 0.09 |
| Congenital anomaly | 0.12 | 0.14 | 0.27 |
| Disability | 0.61 | 1.03 | 1.34 |
| Hospitalization | 12.03 | 27.42 | 30.38 |
| Life-threatening | 0.73 | 4.46 | 1.79 |
| Death | 1.01 | 8.61 | 4.38 |

| Remdesivir |  |  |  |
| --- | --- | --- | --- |
| Not available | 25 | 19.38 | - |
| Other serious (important medical event) | - | 42.71 | 42.23 |
| Required intervention to prevent permanent impairment/damage | 12.5 | 1.02 | 4.3 |
| Congenital anomaly | - | 0.06 | 0.39 |
| Disability | - | 0.71 | 1.27 |
| Hospitalization | 50 | 11.71 | 24.93 |
| Life-threatening | - | 4.72 | 5.87 |
| Death | 12.5 | 19.69 | 21.02 |

The most common uses for hydroxychloroquine were unknown indications (3,760, 15.44%), rheumatoid arthritis (3,288, 13.51%), and coronavirus infection (2,453, 10.01%). The most frequently reported ADRs were pain, arthralgia, headache, and fatigue. A high proportion of the ADR reports (42.68%) resulted in serious medical events, while 22.3% of reports indicated hospitalization. The fatality rate for ADR reports related to hydroxychloroquine was 5.8%. Reports concerning hydroxychloroquine were mainly from the United States (42.19%), Canada (13.54%), and France (7.14%) (Figure 1).

**Figure 1:**
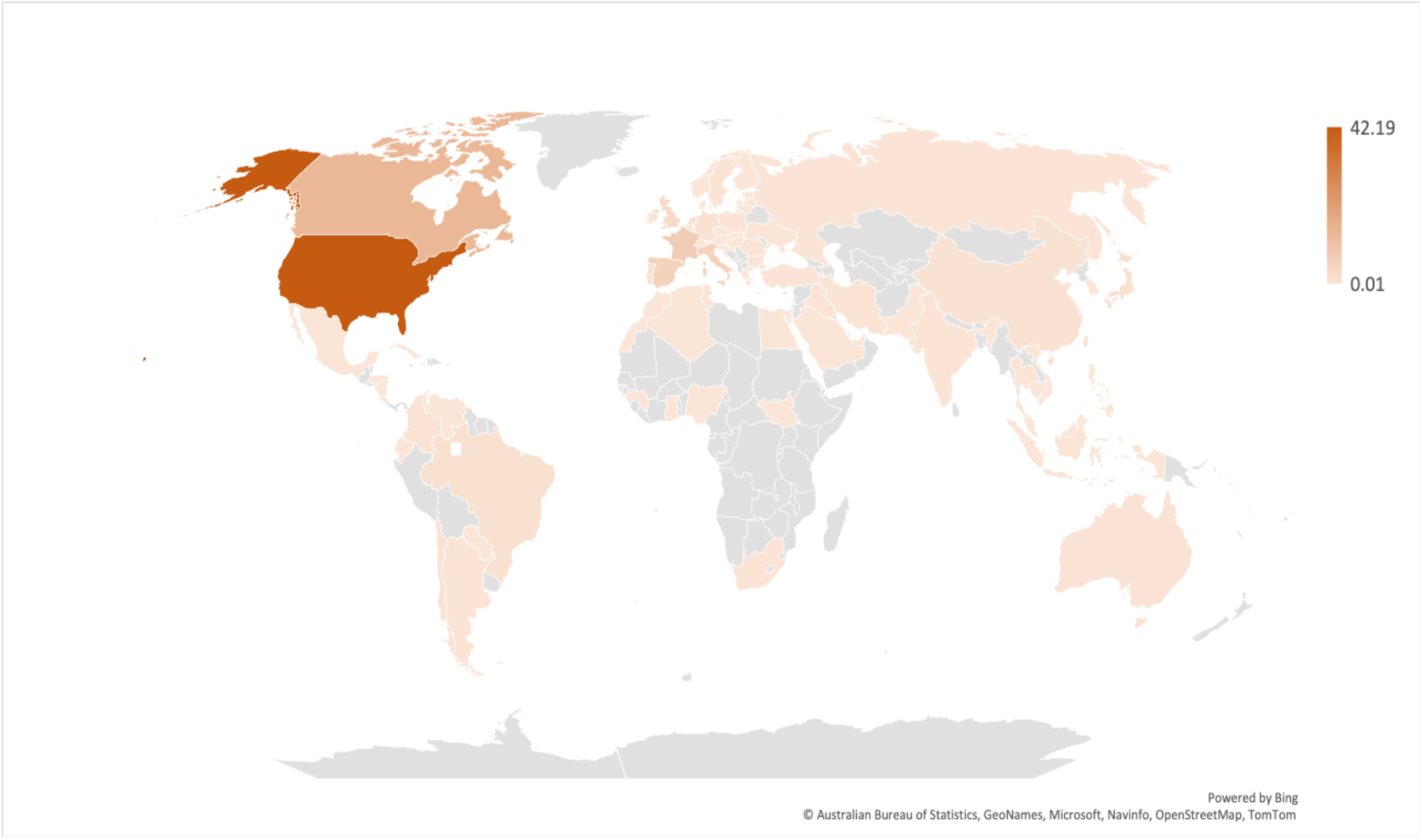
Map showing the percentage of reporting countries for hydroxychloroquine.

More than half of the ADRs related to hydroxychloroquine were reported in 2020 (5,041, 53.3%), compared to 2019 (3,283, 34.7%) and 2021 (1,119, 11.8%). In 2020, hydroxychloroquine was most frequently used for the coronavirus infection (2,428, 18.29%), while unknown indications and rheumatoid arthritis were common during the study period. Before the COVID-19 pandemic, the top three ADRs were arthralgia, pain, and reports of ineffectiveness (Figure 2). Additionally, 969 (29.52%) ADR reports indicated serious medical events, 395 (12.03%) indicated hospitalization, and 33 (1.01%) indicated death (Table 2). However, during the COVID-19 pandemic, the most reported ADRs were QT prolongation (328, 1.48%), and 434 (8.61%) of those cases resulted in death.

**Table 2:** The ROR, PRR, EBGM, and IC for QT prolongation following hydroxychloroquine and fluoroquinolone.

| Drugs | QT prolongation events | Primary suspected exposure | All ADRs | ROR (95% CI) | PRR (95% CI) | IC | EBGM |
| --- | --- | --- | --- | --- | --- | --- | --- |
| Hydroxychloroquine | 58 | 552 | 9443 | 47.28<br>(35.95-62.18) | 42.41<br>(32.25-55.78) | 4.95 | 16.08 |
| Fluoroquinolone | 118 | 5282 | 8179 | 9.32<br>(7.74-11.22) | 9.13<br>(7.59-10.99) | 3.10 | 5.73 |

**Abbreviations:** ADRs: adverse drug reactions, ROR: reporting odds ratio, PRR: proportional reporting ratio, IC: information component, EBGM: empirical Bayesian geometric mean.

**Figure 2:**
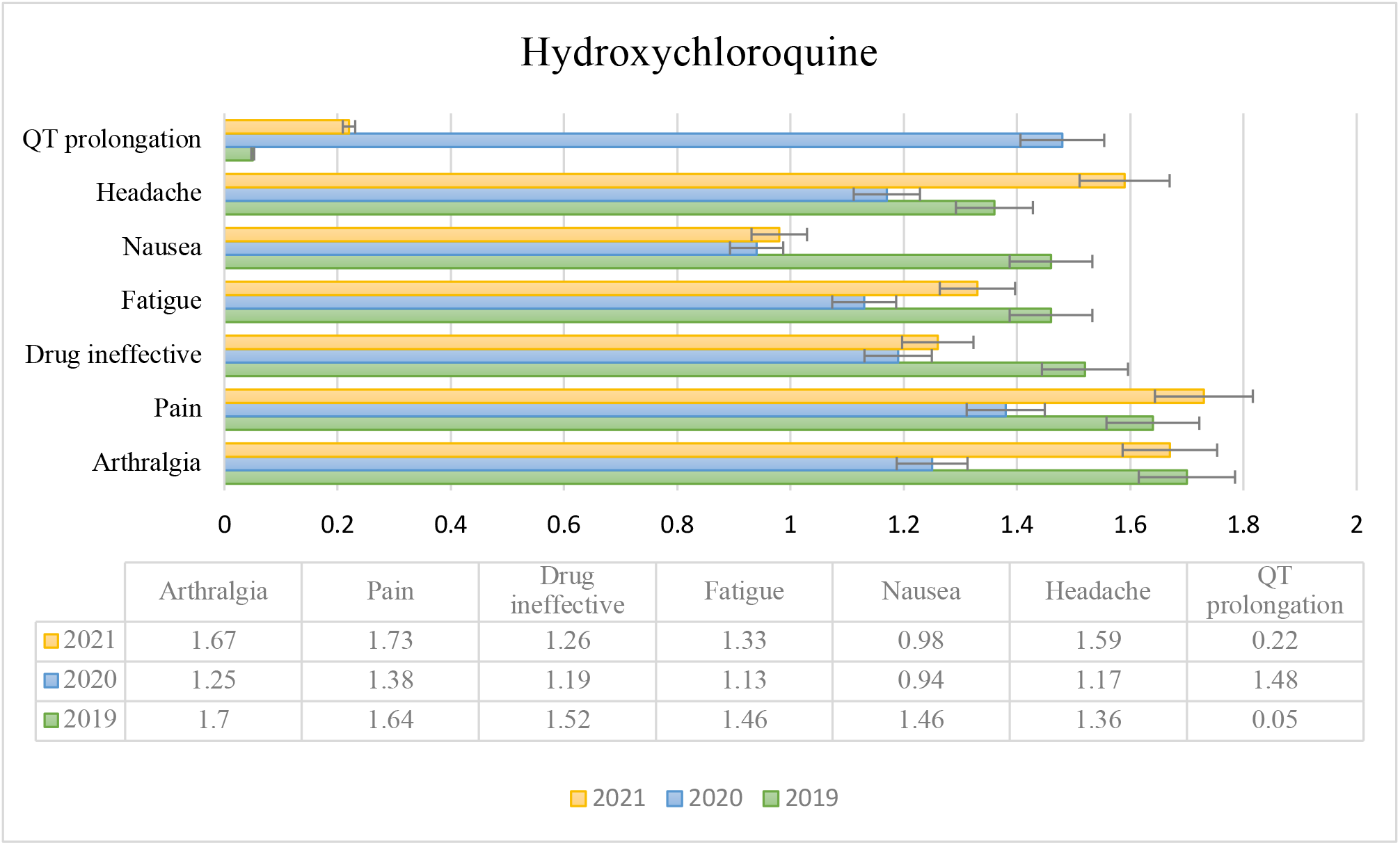
Most frequently ADRs reported for hydroxychloroquine during the study period.

Regarding QT prolongation, all disproportionality analyses showed a statistically significant association with hydroxychloroquine (ROR 47.28 [95% CI 35.95-62.18]; PRR 42.41 [95% CI 32.25-55.78]; EBGM 16.08; IC 4.95). To compare the magnitude of the association of hydroxychloroquine with QT prolongation, we assessed the same signal with fluoroquinolone, and all disproportionality analyses were lower, ROR (9.32 [95% CI 7.74-11.22]; PRR 9.13 [95% CI 7.59-10.99]; EBGM 5.73; IC 3.10) (Table 3).

**Table 3:** The ROR for hepatic and renal events following remdesivir.

| Event | Number of events | Primary suspected exposure | All ADRs | ROR | 95% CI |
| --- | --- | --- | --- | --- | --- |
| Hepatic events | 795 | 5524 | 6673 | 4.81 | 4.46 – 5.19 |
| Renal Events | 365 | 5524 | 6673 | 2.96 | 2.66 – 3.29 |

Remdesivir-related ADR reports involved more males (3,928, 61.13%) and a higher percentage of patients > 65 years (2,949, 44.19%) compared to hydroxychloroquine reports, as described in Table 1. Coronavirus infection was most frequent indication for remdesivir. Elevated liver function test (LFT) (2,427, 14.28%), acute kidney injury (AKI) (857, 5.05%), and bradycardia (502, 2.95%) were the most commonly reported ADRs. The outcome was serious medical events in 42.11% of ADR reports, 14.34% of which required hospitalization, and these events were fatal in 1313 cases. The majority of reports were from United States (5,382, 80.65%), followed by Japan and Italy (298, 4.47% and 150, 2.25%, respectively) (Figure 3).

**Figure 3:**
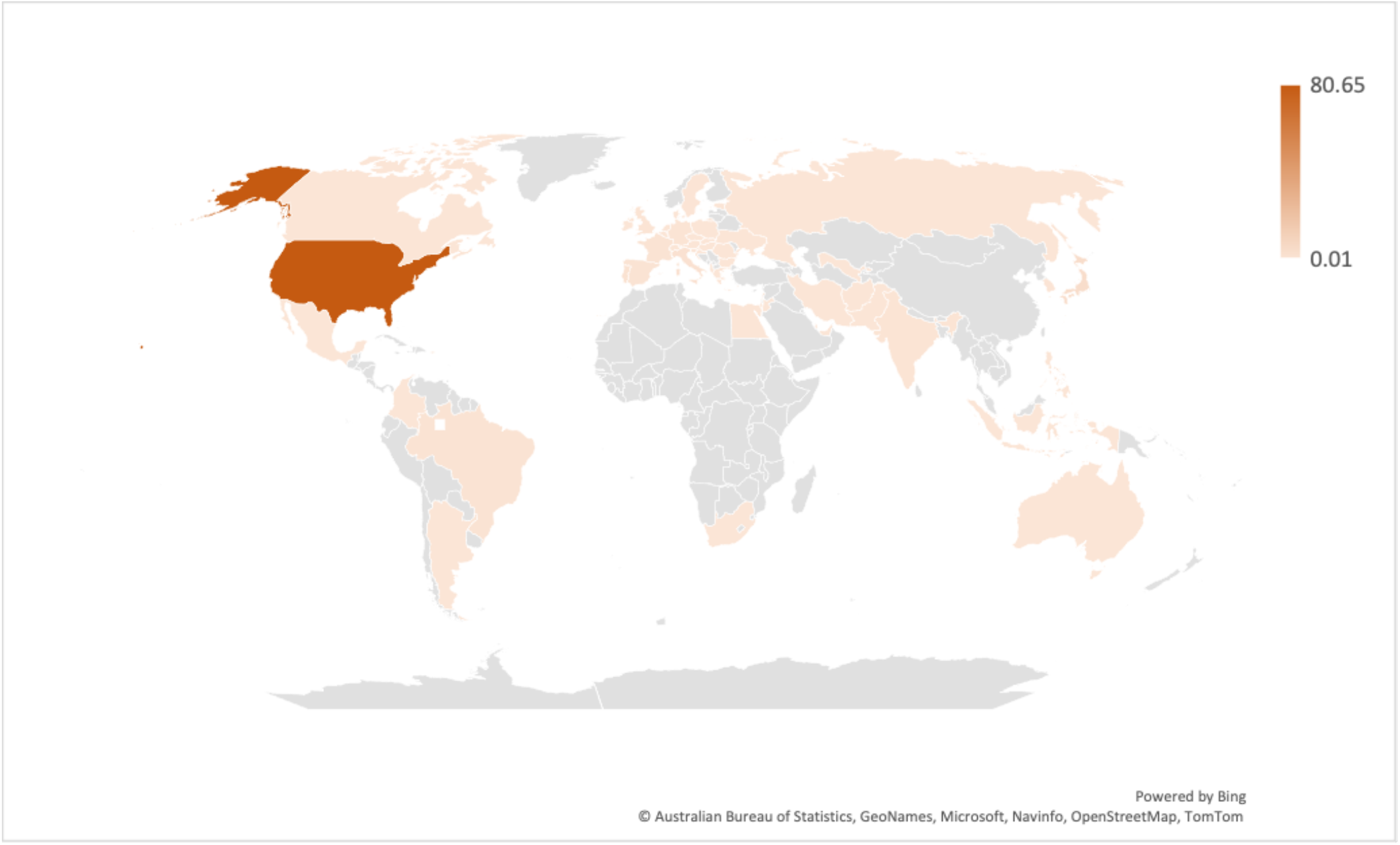
Map showing the percentage of reporting countries for remdesivir.

During 2020, 4,917 ADR reports were associated with the use of remdesivir (84.33% coded as primary suspected drug and 5.72% as concomitant). The main use of remdesivir during the study period was the COVID-19 infection. Elevated LFT (17.26%), AKI (5.95%), and death (2.84%) were the most frequently reported ADRs in 2020 (Figure 4).

**Figure 4:**
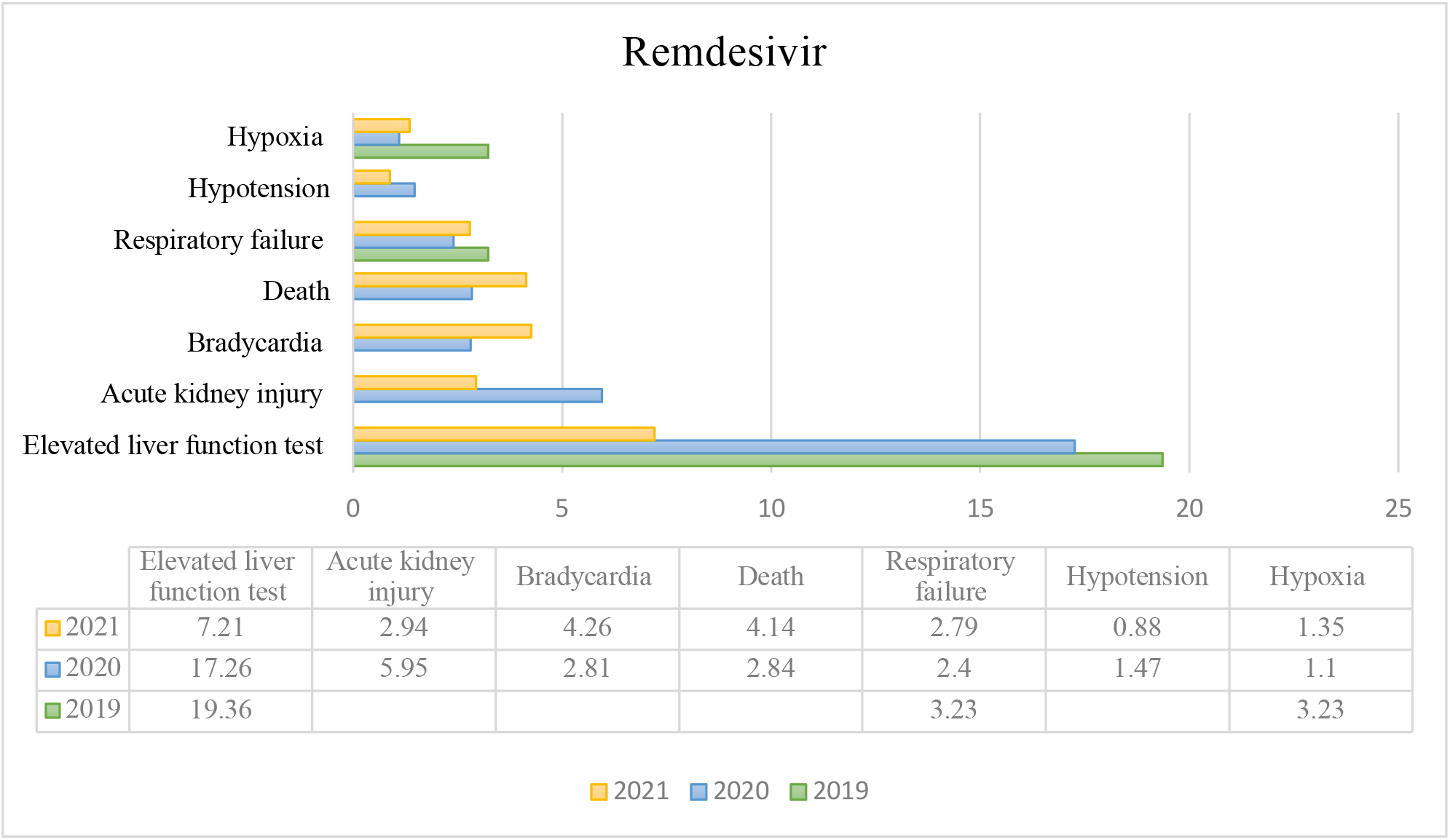
Most frequently ADRs reported for remdesivir during the study period.

We assessed the signals of hepatic and renal adverse events with remdesivir using both ROR and PRR; both were statistically significant (4.81 [95% CI 4.46-5.19] and 2.96 [95% CI 2.66-3.29], respectively) (Table 4).

## Discussion

The sudden emergence and spread of the COVID-19 pandemic caused health care and political leaders to quickly suggest therapies that could potentially treat the disease. Throughout the pandemic, several drugs (including hydroxychloroquine and remdesivir) have been used off-label to treat COVID-19. Later, remdesivir was approved by the FDA. (18) However, there is limited scientific evidence, clinical experience, and safety data associated with the use of these drugs. It is important to note that hydroxychloroquine and remdesivir are reserved for individuals who are infected and are at high risk for developing severe forms of the disease. (19) In this study, we analyzed ADR reports from the FAERS database for hydroxychloroquine and remdesivir from 2019 to 2021.

For ADR reports related to hydroxychloroquine, about 70% of the ADRs were reported in females, and over 30% of the patients of both sexes were over the age of 65 years. The most frequently reported ADRs related to hydroxychloroquine use were QT prolongation, pain, and arthralgia. These reports are consistent with the ADRs reported in other studies. (20-22) For example, the disproportionality analyses in this study showed that hydroxychloroquine was associated with more QT prolongation compared to fluoroquinolone (ROR of 47.38 vs 9.32). A study conducted in Italy found that prolonged QT/long QT syndrome was one of the most frequently reported ADRs (*P* < 0.0008) during the first wave of pandemic. (22) In our study, QT prolongation with hydroxychloroquine use was reported about five times more in 2020, compared to 2019 and 2021. This is because 2020 was the peak of the pandemic; based on our study, about 20% hydroxychloroquine use was related to coronavirus infection. Additionally, the other ADRs reported with hydroxychloroquine (arthralgia, pain, fatigue, nausea, and headache) are considered general symptoms that may have been due to other causes (such as the coronavirus itself), but QT prolongation is a more specific and severe ADR. According to a retrospective observational study, COVID-19 patients had significantly higher risks of torsades de pointes and QT prolongation related to hydroxychloroquine treatment than the non-COVID-19 group (OR 3.10, *P* < 0.001). (23) In addition to concomitant medications, it is worth considering that the risk for QT prolongation is mostly multifactorial, including COVID-19 itself, older age, coexisting cardiovascular disease, and the stage of kidney disease, and each factor’s severity may have a synergistic effect. (24) Furthermore, outcomes related to ADR reports resulted in more hospitalizations and life-threatening conditions during the COVID-19 pandemic compared to 2019. One study showed that 49.3% of ADR reports indicated either hospitalization or the prolongation of existing hospitalization, and 2.9% were related to life-threatening conditions. (22) This is could be explained by increased use of hydroxychloroquine without precautions, which may have led to higher reporting rates.

For ADR reports related to remdesivir, about 60% of reports were in males, and more than 40% of the patients of both sexes were over the age of 65 years. More males experienced ADRs after remdesivir use compared to hydroxychloroquine. We found that elevated LFTs and AKIs related to remdesivir use were the most common ADRs and were statistically significant. In 2019, the most common reported ADRs were elevated LFTs, respiratory failure, and hypoxia. Furthermore, those same ADRs were reported in 2020 and 2021 with additional ADRs reported included AKI, bradycardia, hypotension, and death. The peak of the pandemic occurred in 2020, and remdesivir was approved in October 2020, which explains why many ADRs were reported during that time. A study conducted in 2020 showed that the most frequent ADR associated with remdesivir was hepatic disorder, mainly elevated hepatic enzymes, with ROR 1.94 (95% CI, 1.54-2.45), compared to other drugs prescribed for COVID-19. (25) A prospective study evaluating the compassionate use of remdesivir showed that hepatotoxicity was the most commonly reported ADR, with increases of up to 3-4 grades in transaminase levels, while AKI was the ADR most often resulting in medication discontinuation. (26) A systematic review and meta-analysis suggest that remdesivir is not recommended for patients with elevated LFTs involving alanine aminotransferase and/or aspartate aminotransferase five-fold higher than the upper limit of normal levels and patients with renal impairments either on hemodialysis or with an estimated glomerular filtration rate less than 30 mL/min. (27) This could be because most individuals with a high risk of developing severe cases of COVID-19 are older, with comorbidities such as chronic lung disease, chronic kidney disease, immunocompromised patients, and polypharmacy. (28) In contrast, a systematic review and meta-analysis concluded remdesivir may still effectively reduce the risk of early-stage mortality for hospitalized COVID-19 patients, and it was well tolerated, with an absence of significantly serious adverse events. (29)

This study highlighted different types of ADRs reported to the FDA, rather than focusing on one type. Additionally, our study utilized the FDA’s post-marketing surveillance (FAERS), which is a good resource to assess the use of these medications in the general population. These medications were granted emergency use authorization by the FDA. However, it is always important to undergo a risk-benefit assessment and determine the net favorability of a new drug or a drug used for a new indication. Additionally, this study had a reasonable sample size (about 16,000 reports combined) and had reports from countries around the world, which increases the generalizability.

This study had several limitations. First, we included assessments of two important medications used during the pandemic, but several other therapies used to treat COVID-19 were not studied. There are also several confounding measures that may be missing from the FAERS database, such as disease severity, comorbidities, and concurrent medications. For example, patients who develop QT prolongation while taking hydroxychloroquine may have been on concurrent QT prolongation medications or have precipitating factors that may have increased their risk. However, this event is more likely due to hydroxychloroquine because of the magnitude of the association and increased use of it. In addition, AKI develops in patients with COVID-19, especially if they are in an intensive care unit. (30) This may have confounded our results with the remdesivir group, for example. During the pandemic, spontaneous reporting systems (SRSs) such as FAERS were helpful to detect relationships between specific drugs and adverse drug reactions, but they cannot establish causal relationships. Additionally, SRSs are underutilized and have a chance of inducing reporting bias. (31) Secondly, due to the limitation on prescribing these drugs and negative media attention shortly following the uncovered potential of these drugs, individuals may have not reported their use of these medications.

## Conclusion

Our study highlighted the importance of utilizing scientific evidence when considering any medication for a specific disease, especially with no biological plausible association. This study serves as an important reference to assess the safety of hydroxychloroquine or remdesivir to treat COVID-19. Several serious ADRs were reported with hydroxychloroquine and remdesivir use, as well as hospitalizations and death. It is always important to individualize treatment based on comorbidities, concurrent therapies, and other risk factors to optimize treatments and improve patient outcomes. Additional research and a clinical review should be conducted to assess causality.

## Data Availability

All data produced are available online at FDA website.

https://open.fda.gov/data/faers/

## Notes

### Competing Interest Statement

The authors have declared no competing interest.

### Funding Statement

This study did not receive any funding

